# “We felt like part of a production system”: a qualitative study on women’s experience of mistreatment during childbirth in Switzerland

**DOI:** 10.1101/2021.06.07.21258295

**Authors:** Stephanie Meyer, Eva Cignacco, Settimio Monteverde, Manuel Trachsel, Luigi Raio, Stephan Oelhafen

**Affiliations:** Department of Health Professions, Applied Research & Development in Midwifery, Bern University of Applied Sciences, Bern, Switzerland; Institute of Biomedical Ethics and History of Medicine, University of Zurich, Zurich, Switzerland; Department of Health Professions, School of Nursing, Bern University of Applied Sciences, Bern, Switzerland; Clinical Ethics Unit, University Hospital of Basel and Psychiatric University Clinics Basel, Basel, Switzerland; Department of Obstetrics and Gynaecology, University Hospital of Bern, Switzerland

## Abstract

**Introduction:** Mistreatment during childbirth is an issue of global magnitude that not only violates fundamental human rights but also seriously impacts women’s well-being. The purpose of this study was to gain a better understanding of the phenomenon by exploring the individual experiences of women who reported mistreatment during childbirth in Switzerland.

**Materials and Methods:** This project used a mixed methods approach to investigate women’s experiences of mistreatment during childbirth in general and informal coercion specifically: the present qualitative study expands on the findings from a nationwide online survey on childbirth experience. It combines inductive with theory-guided thematic analysis to study the 7753 comments women wrote in the survey and the subsequent interviews with 11 women who reported being mistreated during childbirth.

**Results:** The women described a wide range of experiences of mistreatment during childbirth in both the survey comments and the interviews. Out of all *survey* participants who wrote at least one comment (n = 3547), 28% described one or more experiences of mistreatment. Six of the seven types of mistreatment listed in Bohren and colleagues’ typology of mistreatment during childbirth were found, the most frequent of which were ineffective communication and lack of informed consent. Five further themes were identified in the *interviews*: informal coercion, risk factors for mistreatment, consequences of mistreatment, examples of good care, and what’s needed to improve maternity care.

**Conclusion:** The findings from this study show that experiences of mistreatment are a reality in Swiss maternity care and give insight into women’s individual experiences as well as how these affect them during and after childbirth. This study emphasises the need to respect women’s autonomy in order to prevent mistreatment and empower women to actively participate in decisions. Both individual and systemic efforts are required to prevent mistreatment and guarantee respectful, dignified, and high-quality maternity care for all.

## Introduction

In recent years, the topic of mistreatment during childbirth has been gaining more and more attention: Women have started to speak out and share their negative experiences of maternity care, which in turn has sparked debate among the public, healthcare systems and the media and has resulted in political and legal change in several countries. A number of Latin American countries for instance have acknowledged obstetric violence as a form of gender-based violence and have passed laws to combat it, following the combined efforts of activists, governmental and non-governmental organisations and researchers, the first of which was Venezuela in 2007 [1]. However, mistreatment during childbirth continues to be a widespread problem around the world, including in high-income countries: Recent studies in Switzerland, Italy, Spain, the Netherlands and the US for instance have shown that a significant proportion of women report that they are mistreated by healthcare professionals (HCP) during childbirth [2-6]. Women have the right to safe, respectful and dignified obstetric care and any form of mistreatment violates fundamental human rights [7-11]. The World Health Organization (WHO) acknowledged the problem in 2015, when it issued a statement for the prevention and elimination of disrespect and abuse during facility-based childbirth in which it called for greater action, dialogue, research and advocacy on the matter [12].

Apart from violating fundamental human rights, mistreatment during childbirth can threaten women’s mental health: negative or traumatic birth experiences are associated with post-traumatic stress disorder (PTSD) [e.g., 13, 14-17]. A meta-analysis on risk factors of postpartum PTSD drew attention to the importance of women’s experiences during birth, reporting an association between PTSD symptoms and low perceived quality of interactions with medical staff [18]. Reed, Sharman and Inglis found that traumatic birth experiences were most often connected to care provider actions and interactions: women reported that care providers prioritised their own agendas over the women’s needs, disregarded women’s own embodied knowledge, used lies and threats to coerce them into complying and violated them [19]. These findings indicate that rather than care providers’ actions themselves, it is the way in which care providers carry these out that is traumatic. Post-partum PTSD can have serious long-term effects for women’s physical, emotional and social well-being and has been shown to severely impair women’s relationships with their partners and their children [20].

One difficulty in gaining a better understanding of and thereby preventing the phenomenon may be the variety of terminology used to describe it: *mistreatment, obstetric violence, disrespect and abuse, dehumanised care*, or *coercion* are often used more or less interchangeably depending on country, context, stakeholders and research methods, although each term emphasises different aspects of the issue [21]. Bohren, Vogel, Hunter, Lutsiv, Makh, Paulo Souza et al. recommend the term *mistreatment of women during childbirth* “as a broader, more inclusive term that better captures the full range of experiences women and healthcare providers have described in the literature” [22]. Their comprehensive, evidence-based typology of mistreatment is the result of a mixed methods systematic review of 65 studies from 34 countries. It encompasses the seven main themes physical abuse, sexual abuse, verbal abuse, stigma and discrimination, failure to meet professional standards of care, poor rapport between women and providers as well as health system conditions and constraints. The typology thereby includes both intentional and unintentional behaviours and acknowledges mistreatment as owing to both individual and structural issues. For these reasons, the present study adopted mistreatment as an umbrella term for violent, disrespectful, abusive, coercive or otherwise poor maternity care.

So far, most studies investigating mistreatment during childbirth have focused on low- and middle-income countries, where the prevalence ranges between 20-77% [e.g., 23, 24-27]. However, recent studies have shown comparable results in high-income countries. In Italy for instance, one in five participants in a nation-wide community-based survey considered themselves victims of obstetric violence [3]. As a result, many decided not to return to the same facility should they become pregnant again or even not to have any more children. Explicit requests for consent before medical procedures were negatively associated with reports of obstetric violence, a finding that highlights the importance of patient autonomy. In a Spanish study, more than a third of the participants indicated having suffered obstetric violence and nearly half reported that they underwent procedures for which they did not provide informed consent [4]. Women who reported obstetric violence were significantly less satisfied with the care they received. Several studies investigating mistreatment during childbirth in high-income countries used the typology by Bohren et al. A secondary qualitative analysis of a phenomenological study on birth trauma with women from New Zealand, the US, Australia and the UK identified six types of mistreatment, the most frequent of which was failure to meet professional standards of care, specifically being neglected and abandoned by HCP [28]. A cross-sectional online survey found that one in six US women experienced at least one type of mistreatment during childbirth, of which being shouted at or scolded by a HCP was the most commonly reported, followed by HCP ignoring women, refusing their requests for help or failing to respond to such requests in a reasonable amount of time [6]. In the Netherlands, van der Pijl, Hollander, van der Linden, Verweij, Holten, Kingma et al. conducted a qualitative social media content analysis of negative or traumatic childbirth experiences shared by women in the #genoeggezwegen campaign (known as #breakthesilence or #rosesrevolution in English) [5]. The most common types of mistreatment were ineffective communication, loss of autonomy and lack of informed consent and confidentiality. In addition to mistreatment, their inductive analysis revealed a further five themes, such as not being taken seriously and not being listened to and short- and long-term consequences, as well as the overarching theme left powerless, which referred to a feeling of powerlessness both during birth and afterwards.

The present project adopted a mixed methods approach to explore the experiences of women who gave birth in Switzerland. The first, quantitative part was an online survey on childbirth experience among over 6000 mothers in Switzerland that focused on *informal coercion* [2]. According to the Swiss Academy of Medical Sciences, medical measures are considered coercive if they are carried out against a patient’s will or despite their opposition or resistance, which highlights the importance of patient autonomy in healthcare [29]. *Formal coercion* is a legally justified restriction of a patient’s autonomy when they lack power of judgment; involuntary committal or compulsory drug treatment for instance are customary measures in psychiatric settings in Switzerland. Seeing as women in labour generally possess power of judgment, less evident or so-called informal methods of coercion may be more likely in maternity care. Informal coercion ranges from relatively covert or subtle forms such as denying women the time or information necessary to reach an autonomous decision to more blatantly coaxing them into complying by means of pressure, intimidation or manipulation or even performing procedures without their consent or despite their resistance [2, 29]. More than a quarter (26.7%) of the women who participated in the survey reported that they had experienced informal coercion during birth [2]. In line with previous research outlined above, experiencing informal coercion was detrimental to women’s birth experience and their mental health: Women who were subjected to informal coercion were less satisfied with their childbirth and were more at risk of post-partum mental health issues. Moreover, migrant women reported higher rates of informal coercion than Swiss women and were more likely to experience post-partum mental health issues.

This article presents the findings from the second, qualitative part of the project, which aimed to expand on the results from the first part and thereby contribute to a better understanding of informal coercion specifically and mistreatment more generally by exploring women’s individual experiences of mistreatment, the aspects of the experience that most affected them during and after birth as well as how they attempted to cope with it. To this end, we first analysed the nearly 8000 comments women left in the online survey and subsequently conducted and analysed interviews with 11 women who reported a negative birth experience.

## Materials and Methods

### Study design

The present research project used a mixed methods approach to assess the prevalence and women’s experience of informal coercion specifically and mistreatment more generally during childbirth in Switzerland. The first, quantitative part was a nationwide, cross-sectional online *survey* on women’s childbirth experience [2]. The survey encompassed questions about their pregnancy and birth in general and focused on informal coercion and the informed consent process regarding any obstetric interventions they underwent specifically. At the end of every section, participants were given the opportunity to provide additional information concerning the previous questions in open-ended text boxes. Due to the vast number as well as the rich content of these comments, it was decided that they should be coded and analysed and included in the second, qualitative part of the project. The key component of this second part was an *interview study* with women who reported a negative experience during childbirth and aimed to expand on the findings from the survey in order to better understand coercion and mistreatment as well as how the women’s experiences affected them during and after birth. One of the authors (StM) constructed the semi-structured interview guide according to the guidelines by Helfferich [30]. Although the main focus of the interviews was informal coercion, it also incorporated questions based on Bohren et al.’s typology of mistreatment during childbirth as well as research on treatment pressure in psychiatric settings [31, 32]. The same author adapted the first draft of the interview guide to feedback provided by the co-authors and subsequently translated it from German to English and French (see S1 Table for English interview guide). The four principal questions addressed women’s overall birth experience, interactions with HCP, decision-making during childbirth and cognitive processing of the birth. At the end, the interviewees were asked a range of demographic questions. Although mistreatment can occur during all stages of pregnancy, labour, birth and the post-partum period, the present study focused the period between the women’s arrival at the birthing facility and the first few hours after birth.

### Recruitment and sample

Out of the 6054 women who completed the *survey* and met the eligibility criteria, more than half (n = 3547) wrote at least one comment in answer to a question potentially relevant to mistreatment, that is, which concerned a specific medical intervention they underwent or informal coercion and the informed consent process in general; the total number of comments was 7753. This constituted the sub-sample and data set for the qualitative analysis of the survey comments. In order to recruit participants for the *interview study*, e-mails were sent out to all the survey participants who had provided their e-mail address for updates on the study (n = 233), inquiring whether they were interested in participating in an interview study on childbirth experience. This e-mail contained a brief and vague description of the goal of the study – i.e., to discover more about women’s individual birth experiences – and a few screening questions, the most important of which was a request to describe their childbirth and any negative experiences they may have had in a few words. Furthermore, we posted a call for participation on our website and sent the same screening questions to the women who answered it. Out of the 50 women who were willing to participate, 11 were chosen for an interview. The inclusion criteria were that they described a negative interaction with HCP during birth, that they had given birth in a facility in Switzerland and that their child was no more than two years old. The women’s age ranged between 29 and 40 years (*M* = 34.8), their children’s between three and 20 months (*M* = 12.3). Seven women were Swiss, two were from neighbouring states (Germany, France) and two were from non-neighbouring states (Poland, Brazil). All women gave birth at a hospital; two by spontaneous vaginal delivery, four by caesarean section and five by vacuum delivery. Regarding their main caregivers during birth, five were primarily treated by doctors and midwives, five by midwives and one by doctors. Eight women were primiparous and three had given birth before. Most women in the sample were highly educated: Eight women had a university degree, one had completed higher technical and vocational training and two had completed basic vocational training. All women apart from one had also participated in the online survey.

### Data collection and management

The *survey* data was collected between August 2019 and January 2020 using the Qualtrics online survey software [33] and subsequently processed in Excel. The *interviews* took place between May and November 2020. Due to the COVID-19 pandemic, it was not possible to conduct the interviews face-to-face. Instead, they were carried using the video conferencing service BlueJeans [34]. The first author (StM) conducted all the interviews; eight in Swiss German, two in English and one in French. Most interviews took roughly half an hour (range: 25 - 77 minutes). Following each interview, the main contents and possible minor adjustments to the interview guide were discussed with another author (SO). The interviews were recorded and subsequently transcribed by one researcher (StM) using the software f4transkript [35]. Information that could allow the direct identification of the interviewees such as their own name or the names of partners, children, HCP, hospitals or places was removed from the transcripts. The interview transcripts were then imported into the ATLAS.ti 8 software for the thematic analysis [36]. One of the authors (StM) translated German and French verbatim quotes into English for the purpose of this publication.

### Ethical considerations

Because this was a national study, all seven regional ethics committees in Switzerland confirmed that by the Swiss Human Research Act, the current study did not require ethical approval (Req-2019-00116). The *survey* participants were asked to confirm that they had understood the study information and to provide their consent on the first page. The *interview* participants were asked to read the written consent form shown through screen sharing and to provide initial verbal consent before the start of the interview. Following the interview, they received the form by e-mail, which they were asked to sign and return. All data were anonymised and treated confidentially.

### Data analysis

Both data sets – i.e., the *survey* comments and the *interview* transcripts – were analysed using thematic analysis [37]. In a first step, one of the authors (StM) coded all 7753 *survey* comments inductively and discussed ambiguous comments, code definitions and examples with another author (SO). As a means of validation and measure of inter-rater agreement, a midwife research intern independently coded one third of the comments. After coding the comments in 1669 survey responses, she received feedback and the codes and definitions were discussed. The inter-rater reliability was calculated based on a validation sample of 533 survey responses. The overall agreement differentiating between women who experienced any form of mistreatment and women who did not was good (Krippendorf’s alpha = .74, 95% bootstrap confidence interval [.66;.82]), although the reliability of the individual codes varied considerably (S2 Table) [38]. The code list generated based on the *survey* comments was not consulted again until the final stage of the qualitative analysis of both data sets to ensure independent, inductive coding and the creation of new codes throughout the *interview* analysis process. In the second part of the qualitative analysis, each interview transcript was first individually coded and then discussed in a group of three to four researchers (StM, SO and one to two midwives per interview). The midwives received only the two to three interviews they were asked to code. They had no insight into the other interviews or the code list based on the previously discussed interviews. Analysis sessions were held for every interview, where the individual codes were examined, compared and discussed and the code list was adapted accordingly. After the final interview analysis session, the two code lists generated during the *survey* and the *interview* analysis were compared to each other and to the typology of mistreatment by Bohren et al. Specifically, the typology of mistreatment was used to structure the data and to assign the codes that matched it to the according themes and sub-themes.

## Results

The two code lists based on the *survey* comments and the *interviews* largely overlapped and many of the codes that referred to mistreatment matched the typology by Bohren et al., even if the exact term often differed (an overview of all themes and codes can be found in S3 Table). Although the original research question focused on informal coercion, the women described a wide range of negative experiences that encompassed not only coercion, but also disrespectful, abusive and physically and verbally violent treatment covered in Bohren et al.’s typology of mistreatment. We found instances of all types of mistreatment according to the typology in the *survey* comments and the *interviews* except for stigma and discrimination, which none of the survey comments in question referenced explicitly enough to justify coding it and was not described in any of the interviews. Physical and/or sexual abuse was only coded in the survey comments since it did not appear in any of the interviews. Seeing as none of the women reported breaches of confidentiality, the according theme was shortened to “lack of informed consent” for the purpose of this paper. In addition to the different types of mistreatment, the inductive coding of the *interviews* allowed the identification of a further five themes, including informal coercion and factors that often co-occurred with mistreatment. The results are divided into two sections: First, the types of mistreatment we identified and their frequencies in the *survey* comments will be presented and subsequently the five themes that emerged in the *interviews* will be described and illustrated with quotes.

### Types and frequencies of mistreatment

Table 3 shows the six out of the seven types of mistreatment and the sub-types according to the typology by Bohren et al. that were found in the *survey* comments and the *interviews*, as well as their frequencies in the *survey* comments. Most comments referred to various types of mistreatment and therefore received multiple codes, which explains why the percentages in Table 3 add up to over 100. Out of the 3547 *survey* participants who wrote at least one comment in answer to a question relevant to mistreatment, 992 (28.0%) described at least one type of mistreatment.

**Table 3.**
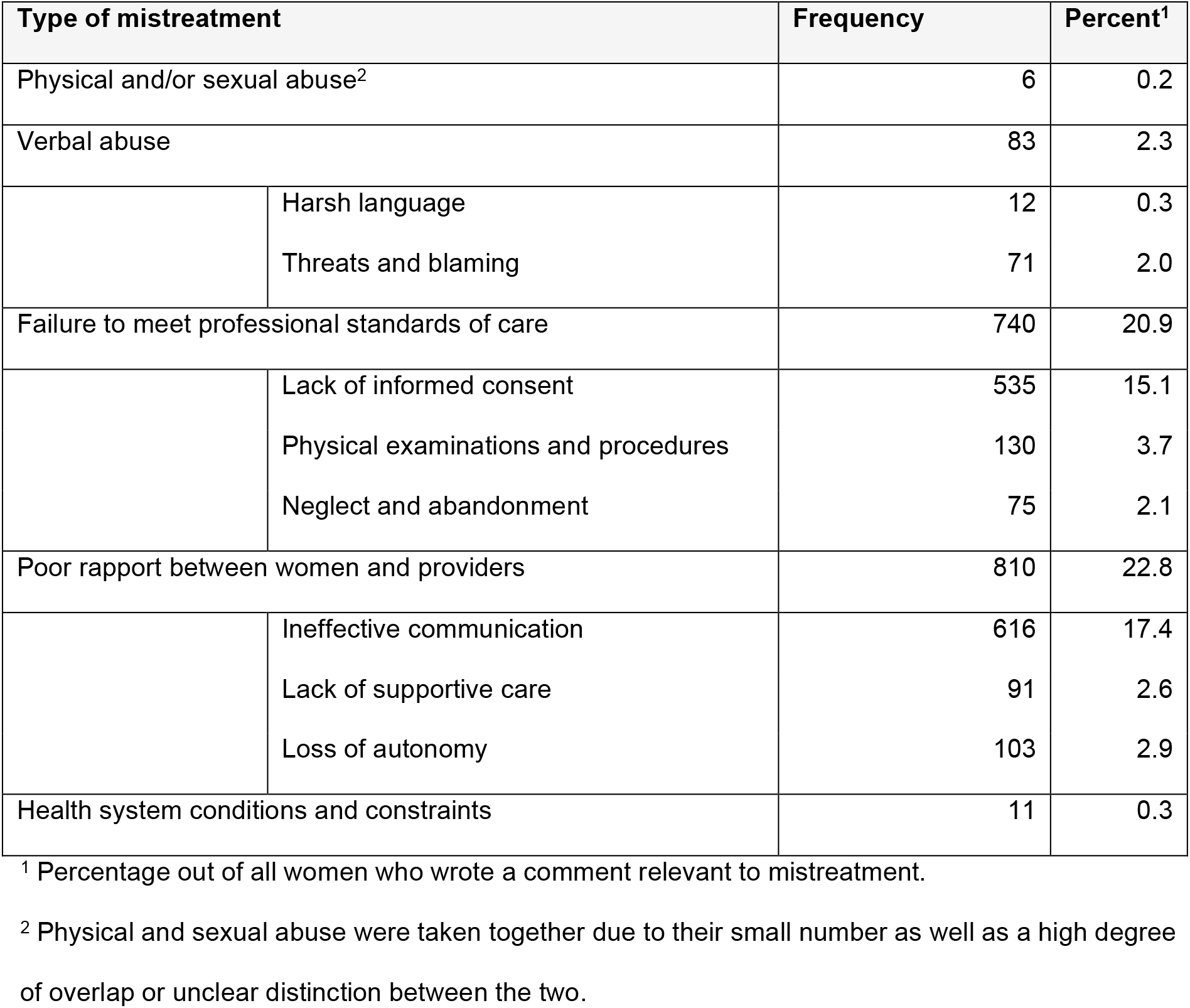
Types of mistreatment and frequencies found in the survey comments.

| Type of mistreatment |  | Frequency | Percent<sup>1</sup> |
| --- | --- | --- | --- |
| Physical and/or sexual abuse<sup>2</sup> |  | 6 | 0.2 |
| Verbal abuse |  | 83 | 2.3 |
|  | Harsh language | 12 | 0.3 |
|  | Threats and blaming | 71 | 2.0 |
| Failure to meet professional standards of care |  | 740 | 20.9 |
|  | Lack of informed consent | 535 | 15.1 |
|  | Physical examinations and procedures | 130 | 3.7 |
|  | Neglect and abandonment | 75 | 2.1 |
| Poor rapport between women and providers |  | 810 | 22.8 |
|  | Ineffective communication | 616 | 17.4 |
|  | Lack of supportive care | 91 | 2.6 |
|  | Loss of autonomy | 103 | 2.9 |
| Health system conditions and constraints |  | 11 | 0.3 |

<sup>1</sup> Percentage out of all women who wrote a comment relevant to mistreatment.
<sup>2</sup> Physical and sexual abuse were taken together due to their small number as well as a high degree of overlap or unclear distinction between the two.

### Interview themes

We identified five themes in the *interviews*: informal coercion, risk factors for mistreatment, consequences of mistreatment, examples of good care and what’s needed to improve maternity care. These five themes and their respective sub-themes are described and illustrated with quotes below.

#### Informal coercion

Informal coercion was mainly characterised by a lack of respect for women and their autonomy, which undermined the informed consent process. The women were often either denied the time and/or information necessary to make an informed choice or not included in decision-making at all. Many of the women described how HCP pressured them to comply, for instance by insisting on an intervention. One woman for example was given morphine during labour despite her repeated opposition.

> *So at first I said, “No, I don’t want anything.” And then it got to a point where it was really tough and I couldn’t think anymore, you know. And then she [the midwife] asked me AGAIN. And then the doctor joined in […] And I think it’s really bad that they didn’t stop the first time I said “No,” […] I wish they’d accepted my first “No.” (Interview 8)*

Several women reported that they were intimidated or threatened with negative consequences for themselves or their baby should they refuse to agree to an intervention, such as caesarean section.

> *I mean, when the doctor came to us and explained that it would be too risky, this means you don’t have choice. So it looked to me like they were deciding and I could not go against it*. […] *So for me it was the right decision in the moment, because of course when you hear, “If you try like this and the umbilical cord gets pressed you can bring problems to your kid,” you don’t wanna risk it. (Interview 5)*

Furthermore, many recounted that they were not taken seriously as individuals, but rather that their wishes, needs, sensations and judgment were dismissed and interventions were carried out despite their concerns. One woman for instance reported that the HCP ignored her when she repeatedly told them the anaesthesia was not effective and proceeded with the caesarean section regardless.

> *Then they asked me again, “Can you feel this, can you feel this.” And, like, each time I said, “Listen, I’m convinced I can feel too much.” I feel like I just knew. […] Now, in retrospect, I think they didn’t really take me seriously when I told them how much I could feel. And that they thought they’d just continue and maybe the effect [of the anaesthetic] would kick in and maybe it would be enough. […] And I honestly feel like I intervened several times […] And then the obstetrician who did the C-section actually started the surgery. (Interview 7)*

#### Risk factors for mistreatment

A number of factors seemed to facilitate mistreatment. Several women talked about how they were not able to think, not able to cope or how they felt insecure or helpless during birth, which left them unable to defend themselves. This in turn often led to procedures being carried out without the women’s consent.

> *I myself didn’t really make any of the decisions. I mean, I don’t remember being asked explicitly. Because by that point I was pretty much delirious, or what do you call that. Even though I hadn’t taken any drugs, but by that point I couldn’t really talk anymore. And then, well, I’d have to ask my husband for details, but I don’t think they asked me, is it ok for you if we do it like this? They just did it [episiotomy and vacuum]. (Interview 3)*

Some women said they were influenced by their partner’s opinion or the inherently unequal caregiver-patient relationship and the resulting questions of trust and responsibility.

> *And then it’s like, if a doctor says to me, “I think we need to,” then yeah. Again, it’s like, how much responsibility do you hand off to them, you know? (Interview 9)*

Furthermore, feeling ill-prepared for birth due to inadequate antenatal classes and prolonged labour often coincided with mistreatment.

> *She said I needed an oxytocin infusion. Like, she thought it was progressing too slowly and that I needed an oxytocin infusion and an epidural. And, like, I don’t know if that had anything to do with the baby or if it was because she decided I couldn’t cope anymore, I don’t know why. (Interview 11)*

#### Consequences of mistreatment

The women reported a wide range of consequences following their experience of mistreatment, which were further divided into four sub-themes: immediate reactions, birth experience, after-effects and coping.

##### Immediate reactions

Women’s immediate reaction to mistreatment depended on their assertiveness and their understanding of the patient-caregiver relationship. More assertive and confident women stood up for themselves by opposing HCP when they felt that their autonomy was not being respected.

> *I really didn’t want a C-section, so I told her obviously, I said, “If there REALLY is a risk for the baby, then you do a C-section of course and I’ll be absolutely fine with that.” I said to her, “But I won’t have a C-section just because it’s too late for you today or because it’s time to get the baby out, like, no.” […] then I said, “ok, you do what you’ve got to do, right, I understand that it’s not your first birth and all.” I said, “But it’s also my belly, so yeah, it’s also up to me whether you open it up or not.” (Interview 10)*

One woman explained how she negotiated with the HCP regarding the handling of her caesarean section.

> *In the end they wanted to give me a general anaesthetic because the epidural wasn’t properly in place. And I said, “No, I don’t want that.” […] they knew I was a surgical nurse […] and he just said to me, “Well if there’s one person who knows what that means it’s you.” And then we made very clear rules, I said they should cut. And the rule or the agreement we had was that, if I really couldn’t take it, that I’d just shout “stop.” (Interview 1)*

Less assertive women or women who seemed to adhere to a more traditional understanding of the roles of patients and caregivers on the other hand often submitted themselves to the HCP. That is, they did not question the HCP’s opinions, gave in to pressure or did not dare to resist. This often coincided with self-doubt and self-deprecation.

> *I think you kind of submit yourself to the team [of HCP] because on the one hand you trust they know what they’re doing and that they’ll do it well and also because you, like, don’t want to be the one who interferes. (Interview 7)*
>
> *I mean, we had no idea why it was going so fast and we also didn’t speak up, I think maybe we could also have said it again. But when I said it was going too fast, when I told the midwife, “Hey, I asked you for a few minutes,” and she said, “No, we need to start,” then I am not the one confronting too much. (Interview 5)*

##### Birth experience

Looking back on their birth, most women who experienced mistreatment expressed negative feelings regarding the birth as a whole or specific parts of it. Asked about how mistreatment made them feel during birth, some women described a loss of control or autonomy that left them in a state of distress.

> *And we felt like, really like part of a production system, you know. We felt in a rush, like everything has to be in a rush, and it’s ta ta ta ta ta, and we really felt extremely bad with this, it was terrible for us. (Interview 5)*

One woman recounted how she felt robbed of a positive birth experience due to the various types of mistreatment she suffered at the hands of her main caregiver.

> *Someone once asked me, “Are you looking forward to the birth?” And that wasn’t the case for me, but I think you can still have a good birth experience. And she [the midwife] took that away from me […] I’ve never felt so completely and utterly at someone’s mercy before. (Interview 11)*

##### After-effects

Most women who reported being mistreated struggled with coming to terms with their experience and felt sad or frustrated following their birth. These feelings often persisted for a long time after birth or even had lasting effects. While some were left with open questions and uncertainties they needed to clarify with HCP, others avoided thinking or talking about their experience altogether. Many were troubled by “what if” thoughts, that is, they continued to go over scenarios of how their birth could have unfolded differently.

> *That’s why I had to go to my gynaecologist and talk to her, because I was feeling extremely frustrated. And I always had in my head, on the one hand we made a good decision thinking of the risk, on the other hand I always had in my head, and if? And if we would have waited a bit more? You know, I asked for some time to talk to my husband, I was not asking for time to see if the baby would turn, but maybe five or ten minutes more and my baby would have turned and the story would have been completely different. And I think I will always carry this doubt with me, but I have to carry the point that, sorry, I get a bit emotional, but I have to carry the point that the decision was made based on risk and what was safest for us. (Interview 5)*

Many also struggled with regrets or blamed themselves for what they experienced.

> *It wasn’t until later that I felt, like, guilty about how it all turned out. That it was my fault […] in the first few weeks I really struggled with that. I was just sad afterwards because I didn’t understand what happened there at all. And I had no control over it. And feeling kind of like guilty that I did it all wrong and that I could have done better and that it would’ve turned out differently. (Interview 6)*

Some women described how their experience of mistreatment affected or changed their feelings, attitudes or intentions regarding a potential future pregnancy and birth.

> *When I, like, think about having a second child, that’s something I tell myself I don’t really want to experience again. I’m scared of that. And I think if I got pregnant again, then I’d probably be really preoccupied with that fear in the months before the birth. (Interview 6)*

##### Coping

The women often pointed out how it took time for them come to terms with or let go of what happened to them. In terms of processing their experience, most women found talking about their birth helpful and important, be it with the HCP who were involved, with other (mental) health providers or with their family and friends.

> *And then a nurse came and she, like, asked me why I wasn’t feeling so good. And I kind of, like, said I was still thinking about the birth and she really listened to me. That’s, like, the only time during my whole stay at the hospital where I felt like someone took time for me. And I kind of discussed everything with her and she said, like, she could write it down and then I could also talk to the midwives about it. (Interview 9)*
>
> *I think the conversation with the midwife was most helpful to me and definitely also talking to other people […] Like, when someone asked me what it was like, I mean I was always pretty honest. And I was able to discuss it with friends of mine who’ve already given birth themselves. (Interview 1)*

Extenuating their mistreatment seemed to be another common form of coping, seeing as all interviewees diminished their experience or exonerated the individual HCP or the facilities who mistreated them. Instead, they blamed themselves or responded with understanding and searched for justifications and external or situational explanations.

> *I don’t think she, like, did a bad job on the whole. I think maybe she’s also just a victim of the hospital environment, where those drugs are just available, you know […] And I’m not angry or anything, not at all, I just wish it had been different. (Interview 8)*

Some women also compared themselves to others whose experience they judged as worse than their own or focused on positive aspects of their birth or on the fact that their baby was born healthy.

> *And I don’t know, in retrospect I sometimes think maybe she wouldn’t have needed to go to the paediatric hospital and it all would’ve been much more relaxed if it hadn’t been such a knee-jerk reaction. But, like, I can’t say, that’s not my area of expertise, like, I wouldn’t want to blame anyone. The main thing is she’s healthy now. (Interview 11)*

#### Examples of good care

Some women also recounted positive experiences and interactions with HCP during childbirth, such as HCP treating them with empathy, communicating openly, respecting boundaries and patient autonomy as well as enabling or even encouraging them to make their own informed decision. Examples of good care also included HCP giving women time to come to terms with a decision even after it had essentially been made.

> *I mean, they at least let me feel like it was my decision. And I know there was no way around the C-section, but it was up to me to decide when it happened. It was up to me whether I needed another half an hour or whether I wanted to do it straightaway, I was even asked if I needed more time to consider. (Interview 1)*

HCP who treated them with respect and took the time to explain procedures left a good impression on women.

> *They even showed me what [the vacuum] looked like and how it worked first. And said, like, that it was possible there’d be a little bump, but, “This is what it looks like, this is how it works and now we hope we can help get [the baby] out.” So, yeah, I think those two doctors were very good and the midwife too. (Interview 2)*

#### What’s needed to improve maternity care

The last theme encompasses women’s own wishes and suggestions for improving maternity care; both what they would have wanted during their own childbirth as well as general improvement suggestions. Almost all women stressed the need for HCP to treat them with respect and empathy, to provide more support and help and to take more time for them.

> *So I just get the impression that giving birth in general and maybe the whole time after that, especially the first few days, is such an exceptional situation in a person’s life and I sometimes just think that people aren’t sensitive enough or maybe don’t have enough time to be sensitive. I think it has a lot to do with the time factor, I mean with those case rates, that you almost feel as if you’re being kicked out. Or like, everything is timed at the hospital, so the individual sometimes comes up short. Yeah or maybe that it really stands or falls on people taking time for you now and again despite time constraints. Like, it leaves a good impression if someone really takes time for you. (Interview 9)*

Several women indicated that they did not understand the interventions they underwent or the reasons for them and that they would have required more orientation. Some also recounted that they felt rushed and wished for more time to consider and process information.

> *I think maybe in the first talk having more time to talk. Because it was ok, he came and he explained everything, he was very nice and he explained it very nicely, but then that was it, and then of course he asks you if you have questions, but I think in your head you are still, you know, processing all the information. And as everything was too fast, I didn’t have time to even think […] I didn’t have time to think about anything else and say, “I’d like to talk to the doctor more.” (Interview 5)*

The interviews made it clear that women’s relationship with the HCP was a key aspect of their birth experience, seeing as they emphasised the need for a main caregiver they knew they could trust and supported them.

> *And out of all the people involved, I must say there was always someone I could trust or felt comfortable with and I think that’s the most important thing. I don’t think it’s necessary for everyone to be like that, but definitely to have one or two people where you feel like, yeah, this is someone I can trust or who makes me feel good. (Interview 2)*

## Discussion

The present study aimed to expand on the results from a nationwide survey on childbirth experience and thereby contribute to a better understanding of mistreatment during childbirth and how it affects women during and after birth. To this end, we analysed the comments collected in the *survey* and the *interviews* that were conducted with women who reported a negative experience during childbirth. Our findings indicate that women are subjected to various types of mistreatment during childbirth in Switzerland, which has a negative impact on their birth experience and their well-being. In addition to mistreatment, we identified informal coercion as a specific type of mistreatment, risk factors for and consequences of mistreatment, examples of good care and women’s perspectives on what is needed to improve maternity care in the interviews.

The quantitative analysis of the *survey* comments showed that the number of women who described mistreatment in a comment – i.e., 28.0% of all women who wrote a comment – matched the 26.7% of women who reported informal coercion in the previously published quantitative part of the project [2]. Although the initial research question focused on informal coercion, women described various types of mistreatment according to Bohren et al.’s typology. The two most common types of mistreatment found in the survey comments were ineffective communication, which was coded in 17.4% of the comments, and lack of informed consent, which 15.1% of the comments referred to.

These were also two of the three most reported types of mistreatment in van der Pijl et al.’s study in the Netherlands [5]. It seems many women do not receive sufficient information or orientation during childbirth and are not respected as individuals, which leaves them feeling insecure or passive and undermines the informed consent process. An interesting finding was the difference in language used to describe experiences of mistreatment between the survey comments and the interviews. While the survey participants used more explicit and more negative words to describe their experience, the interviewees were more cautious and chose less explicit words. It may be easier to report such an intimate and distressing experience anonymously in an online survey than to describe it in an interview, which underscores the importance of combining the two approaches. A study with postpartum women showed that they disclosed significantly more sensitive information such as intimate partner violence when they were guaranteed anonymity compared to confidentiality of their responses [39]. The fact that many of the survey participants and all interviewees described several types of mistreatment suggests that mistreatment is rarely a single, isolated event, but rather that certain types of mistreatment frequently co-occur or may even give rise to others. For example, failure to respect a woman’s autonomy and to communicate effectively undermines the informed consent process, which in turn can result in procedures being carried out against her will. The overlap and relationship between different types of mistreatment is an avenue worth exploring in future research.

In addition to mistreatment, a further five themes emerged through in the *interviews*: informal coercion, potential risk factors for mistreatment, consequences of mistreatment, examples of good care and what’s needed to improve maternity care. These themes can precede, co-occur with or follow mistreatment and thus provide a fuller picture and a broader understanding of the phenomenon.

Informal coercion was identified as a central theme in the interviews and was characterised by a lack of respect for women’s autonomy. This finding highlights the importance of enabling and encouraging women to actively participate in decisions regarding their care. This can be achieved through *shared decision-making* (SDM), which promotes open, honest and respectful communication and involves patients and HCP collaborating to make health care decisions that are based upon the best available evidence and patients’ preferences, values and needs [40]. SDM is therefore argued to be “the pinnacle of patient-centred care” [41]. In order for HCP to put SDM into practice however, they must subscribe to the guiding ethical principle that individual self-determination is a desirable goal and support patients to achieve this goal [42]. Although SDM is beneficial to patients, HCP and health care facilities and systems alike, a number of barriers still prevent it from being the norm in maternity care, which include “existing cultural norms of ‘the doctor knows best’ and ‘patient acquiescence’” [43]. Begley, Daly, Panda and Begley argue that other factors such as concerns over litigation, private insurance that pays more to the obstetrician if more interventions are carried out, constraining guidelines and management policies, HCP’s level of confidence and skills and HCP making decisions based on their personal convenience are most likely additional barriers in the way of SDM [43]. Therefore, structural and systemic change is needed to foster acceptance of a culture of SDM and thereby implement it as the norm in maternity care.

A number of factors were discerned that often occurred in the context of mistreatment and therefore may contribute to, facilitate or exacerbate it. These risk factors pertained to the woman (e.g., “not being able to think”), to interactions with the HCP or her partner (e.g., the power imbalance between HCP and patients) or to the situation (e.g., prolonged labour). While this finding is consistent with studies that have demonstrated effects of socio-demographic factors such as nationality or race on mistreatment, the present study does not allow any causal interpretations and future research should investigate the link between these potential risk factors and mistreatment [2, 6].

Many of the women reported negative consequences of mistreatment that affected them both in the short and/or long term. This was the case for both comparably mild and more severe experiences of mistreatment. This finding corresponds with van der Pijl et al.’s findings, who identified a theme by the same name in their inductive analysis, which most often referred to emotional consequences that tormented some women even years later [5]. In the present study, more assertive women reacted to mistreatment with resistance and defended their autonomy during childbirth, while less assertive women or women who seemed to have a more traditional understanding of the patient-caregiver relationship submitted themselves to the HCP. This finding emphasises the need to respect women’s autonomy and to pay particular attention when treating less assertive or primiparous women. Experiencing mistreatment resulted in a negative appraisal of the whole birth experience or parts of it. This ties in with previous studies that found negative effects of mistreatment on women’s satisfaction with childbirth [2, 4]. For some women, mistreatment resulted in a change in attitudes or intentions regarding (future) childbirth. In line with the previously referenced Italian study, certain women decided not to return to the same facility if they became pregnant again or even not to have any more children [3]. Many dealt with after-effects such as feelings of sadness and frustration, guilt and regret, self-blame, flashbacks, obsessing over “what if” scenarios and how their experience could have been different or avoiding memories of their birth. Byrne, Egan, Mac Neela and Sarma, who explored the subjective experience of birth trauma, found that coping through detachment, which encompasses avoidance and dissociation, was a common response to a traumatic birth experience [44]. While the present study did not assess mental health indicators, the previously described after-effects are established symptoms of PTSD. Furthermore, some women reported they had been diagnosed with PPD and/or that they were currently receiving psychological treatment to come to terms with their birth. These results tie in with previous research that has confirmed a link between a negative or traumatic birth experience and post-partum mental health issues [13-19].

The women described different ways of coping with their experience of mistreatment, such as discussing it with other people or extenuating it. Asked about what they found most helpful in processing their experience, most women stressed the importance of talking about it, be it with the HCP involved, with other (mental) health providers such as nurses or therapists or with family and friends. Women who had avoided thinking or talking about their mistreatment up until the interview exhibited clear signs of not having fully processed their experience and not being at peace with it. Although there is limited evidence for the effectiveness of debriefing interventions in preventing postpartum mental health issues, several reviews nonetheless support our finding that women generally find talking about their experience and being listened to beneficial [45-47]. One of the most frequently used codes was extenuation, seeing as all women diminished or excused their mistreatment. This ties in with Ayers, who found that women who self-reported a traumatic birth, regardless of whether they had PTSD symptoms or not, engaged in various thought processes to help cognitively process their experience such as retrospectively taking a fatalistic view of birth or focusing on the inevitability of labour [48]. Our findings concerning the consequences of and women’s coping with mistreatment during childbirth give evidence to the serious and long-lasting impact it can have on women’s well-being and to the benefits of sharing their experience. Furthermore, they indicate the need for sensitive after-care for all mothers to support them in processing their birth and ideally detect and treat postpartum mental health issues.

Although the present project centred around mistreatment, some women also described examples of good care and positive experiences and interactions with HCP during birth. These accounts correspond with aspects of Shakibazadeh, Namadian, Bohren, Vogel, Rashidian, Nogueira Pileggi et al.’s typology of respectful maternity care (RMC), which is the result of a qualitative evidence synthesis of 67 studies from 32 countries and encompasses twelve domains, such as prospective provision of information and seeking informed consent, engaging with effective communication and respecting women’s choices [49]. RMC refers to “care organized for and provided to all women in a manner that maintains their dignity, privacy and confidentiality, ensures freedom from harm and mistreatment, and enables informed choice and continuous support during labour and childbirth” and is recommended by the WHO [50]. So far, research on the effectiveness of such policies is sparse. A systematic review of five studies undertaken in four African countries found moderate evidence that multi-component RMC policies reduced women’s experiences of disrespect and abuse overall [51]. However, there was less evidence for an effect on individual types of mistreatment. Moreover, the most successful elements as well as the sustainability of such policies remain unclear. The fact that some women in our study experienced elements of both mistreatment and respectful care confirms that the two do not necessarily exclude each other and that RMC is more complex than the mere absence of mistreatment [21, 52]. This in turn suggests that preventing mistreatment and implementing RMC may require simultaneous yet different types of interventions [21, 52]. Therefore, researchers and health care providers alike must strive to prevent mistreatment as well as implement RMC in order to ensure respectful and dignified care for all – before, during and after childbirth.

The last theme encompassed women’s perspectives on what is needed to improve maternity care. Making women’s voices heard was the aim of the *What Women Want* global advocacy campaign, which seeks to improve quality maternal and reproductive healthcare and strengthen health systems. The campaigners asked 1.2 million women in 114 countries about their priorities regarding maternal and reproductive health services and their top demand was respectful and dignified care [53]. This corresponds with our finding that most women stressed the need for more sensitive care and for HCP to treat them as individuals, which was particularly well illustrated in a quote by one interviewee who emphasised the importance of HCP acknowledging birth as an exceptional situation and experience and taking time for women despite time constraints. Therefore, in order to truly attain high quality women-centred maternity care, it is essential to listen to women individually and collectively and to incorporate their insights into healthcare policies and guidelines.

Taken together, our findings highlight how multifaceted and multi-layered mistreatment during childbirth is. While the interpersonal level of the caregiver-patient relationship may be the most obvious and interventions directed at HCP are essential, the systemic and structural level must not be ignored. However, while systemic factors can provide contextual explanations for mistreatment, they can never serve as justifications.

### Strengths and limitations

One of the primary strengths of the study is its design, which integrated two different data sets: the comments written in a largescale, nationwide online survey on childbirth experience and the interviews conducted with women who reported being mistreated during childbirth. This allowed both quantitative conclusions regarding the frequency of different types of mistreatment as well as qualitative conclusions concerning women’s individual experiences of mistreatment and how these affect them during and after childbirth. Furthermore, the qualitative analysis combined inductive with theory-guided thematic analysis. On the one hand, this approach confirmed the applicability of Bohren et al.’s typology of mistreatment in a Swiss context. On the other, it allowed a further five themes to be identified that contribute to a fuller picture and a broader understanding of the phenomenon.

The limitations of this study include the self-selection of participants, both for the survey and for the interviews. In the *survey*, women could choose to provide further information about their birth in open-ended text boxes at the end of every section, however, this was not mandatory. As for the *interviews*, the previously referenced tendency to disclose more sensitive information in an anonymous setting may also have played into the relatively low response rate of 21% to the recruitment e-mail, especially considering that the women it was sent to had all already filled in the questionnaire [39]. Women who chose to participate in the interview study are likely to differ from women who did not. For instance, women who had a particularly traumatic experience and are coping through detachment may shy away from sharing their story in an interview.

Another important limitation of the study is the lack of diversity in the interview sample, considering that all interviewees were white heterosexual cisgender women who were Swiss nationals or residents and mostly highly educated. Future research should include marginalised populations such as the LGBTIQ+ community and refugees and asylum-seekers. Seeing as marginalised groups are more vulnerable to stigma and discrimination in general, they are also likely to be more at risk of mistreatment in maternity care [6, 54]. The previously referenced US study for instance found that Black, indigenous and women of colour experienced significantly higher rates of mistreatment [6]. Furthermore, the sample only included women living in the German speaking part of Switzerland, since no one from the French speaking part met the eligibility criteria and no one from the Italian speaking part replied to the recruitment e-mail.

Lastly, it seems important to point out that the interviews took place during the COVID-19 pandemic. The effects of this context in itself and the fact that the interviews took place by video call as a consequence are unclear. Although many aspects of our life have shifted to the digital sphere and we have become accustomed to life online over the course of the pandemic, some important aspects that are inherent to face-to-face interactions such as nonverbal signals and body language cannot be conveyed in a video call. Furthermore, meeting in person provides the opportunity for building trust through small talk prior to an interview. It is therefore possible that face-to-face interviews would have encouraged women to share more intimate details. Despite these limitations, the study shows that mistreatment is a reality in Swiss maternity care and affects women both during and after birth and introduces avenues worthy of further investigation.

## Conclusions

Research has shown that mistreatment during childbirth is an issue of global magnitude, which was acknowledged by both the WHO and the UN. The results from the present project confirm and extend previous findings by demonstrating that women in Switzerland face various types of mistreatment during childbirth. Furthermore, this study sheds light on women’s individual experiences of mistreatment and how these affect them during and after childbirth. It pinpoints informal coercion as a specific type of mistreatment, identifies risk factors for and consequences of mistreatment as well as examples of good care and amplifies women’s views on what is needed to improve maternity care. The findings from this study thus contribute to a better understanding of mistreatment during childbirth and highlight the need for systemic efforts to increase HCP’s awareness of the issue and to enable and empower women to actively participate in decision-making in order to achieve respectful, dignified and high-quality maternity care for all.

## Data Availability

The data cannot be shared publicly because they contain sensitive information and could allow individuals to be identified. When consenting to participate in the study, the women were guaranteed that anonymity and confidentiality would be strictly maintained and that only the researchers involved in the project would have access to the data collected. Data may be made available upon reasonable request. For requests please contact.

## Acknowledgements

First and foremost, we are incredibly grateful to the women who took the time to share their birth experience with us. We would like to thank Lea Bucher, Anja Hurni, Raquel Mühlheim, Cecilia Gebhart and Rahel Messmer for their valuable contribution to the analysis of the interviews.

## Author contributions

**Conceptualisation and design**: all authors.

**Project administration**: Stephanie Meyer, Stephan Oelhafen.

**Methodology**: Stephanie Meyer, Stephan Oelhafen.

**Data collection**: Stephanie Meyer, Stephan Oelhafen.

**Data analysis**: Stephanie Meyer, Stephan Oelhafen, Eva Cignacco.

**Preparation of the manuscript**: Stephanie Meyer.

**Review of manuscript draft and approval of the final manuscript**: all authors.

## Supporting information

**S1 Table. English interview guide**.

**S2 Table. Inter-rater reliability of survey coding**.

**S3 Table. Overview of all themes and codes**.

