## Supplementary material for "“We felt like part of a production system”: a qualitative study on women’s experience of mistreatment during childbirth in Switzerland": S1 Table

**S1 Table. English interview guide.** ^1^ Only asked if indicated.

| **Key question** | **Checklist and prompts for further questions** | **Directive questions** |
| --- | --- | --- |
| Birth experience | | |
| I would like to begin with your birth experience. Can you tell me about how your birth unfolded from the moment you arrived at the facility? | - Did the birth go more or less as planned or as you had envisioned it?   - Why yes/no? - How did you feel during the birth? - Was there anything that didn’t go so well? - Was there a specific moment, when the atmosphere changed, you no longer felt comfortable or the situation got out of hand?^1^ | - Nonverbal - Can you tell me more about that? - What happened next? - What about …? |
| Interactions with healthcare professionals | | |
| My next question concerns your interactions with the healthcare professionals during birth (midwives, doctors, possibly students). What did you think of the care provided by the healthcare professionals? | - Did you feel respected? - Did you feel taken seriously? (Regarding your feelings, wishes, needs) - Did you feel supported at all times? - What did you think of the communication and collaboration among the different healthcare professionals? | - Nonverbal - Can you tell me more about that? - What happened next? - What about …? |
| Decision-making | | |
| Next, I would like to know more about the decision-making process during your birth. Please think of a situation where an important decision had to be made. How did that decision come about? | - Did the healthcare professionals include you in the decision-making process? - Was it important to you to have an active say during your birth? - Did the healthcare professionals always inform you of their plans and of every step of their procedure? - Could you always understand what they were doing and why they were doing it? - Did you have enough time to consider the decision? - Did the healthcare professionals always ask for your consent? - Who made the final decision? - How did you weigh the options, what were your thoughts and feelings regarding the decision? - Did you always trust the healthcare professionals? - Did you and the healthcare professionals ever disagree on anything?   - What did you disagree on?^1^   - Did you express your opinion?^1^     - If yes: How did the healthcare professionals react?^1^     - If no: Why not?^1^   - Can you tell me more about the communication with the healthcare professionals?^1^     - What were their arguments?^1^     - Were there different consequences depending on which decision was made?^1^   - Who convinced who? Or did you find a third solution?^1^ | - Nonverbal - Can you tell me more about that? - What happened next? - What about …? |
| Processing the birth | | |
| Lastly, I would like to know how you look back on your birth today. | - Is there anything about your birth that is still on your mind today? - What was most helpful to you in processing your birth experience? - Did you have the opportunity to discuss the birth with any of the healthcare professionals involved after the birth?   - If yes:     - Was it helpful?^1^     - Why yes/no?^1^   - If no:     - What was missing?^1^     - What would have helped you?^1^     - Why not?^1^     - Did they offer you the opportunity?^1^ - Did you see the need? | - Nonverbal - Can you tell me more about that? - What happened next? - What about …? |
| End | | |
| We’ve reached the end of the interview. Is there anything we haven’t talked about that you’d like to add? |  |  |
