## Supplementary material for "“We felt like part of a production system”: a qualitative study on women’s experience of mistreatment during childbirth in Switzerland": S2 Table

**S2 Table. Inter-rater reliability of individual survey codes**. Krippendorf’s alpha was only calculated for codes with at least 30 instances.

| **Code** | **Krippendorf's alpha [95% confidence intervals]** |
| --- | --- |
| **Mistreatment (overall)** | **0.74 [0.66;0.82]** |
| Not taken seriously | 0.67 [0.46;0.83] |
| Lack of consent | 0.69 [0.50;0.84] |
| Pressure | 0.45 [0.21;0.67] |
| Lack of information | 0.52 [0.19;0.77] |
| Lack of respect for wishes/feelings/needs | 0.66 [0.36;0.87] |
| Degradation | 0.28 [-0.01;0.67] |
| Restriction of freedom | 0.62 [0.25;0.86] |
| Unprofessional conduct | 0.62 [0.22;0.86] |
| Median [range] | 0.62 [0.28-0.69] |
