## Supplementary material for "“We felt like part of a production system”: a qualitative study on women’s experience of mistreatment during childbirth in Switzerland": S3 Table

**S3 Table. Overview of all themes and codes.** ^1^ The codes were translated from German to English by one author (StM). ^2^ Miscellaneous codes were not included in the analyses because they were not relevant for the present research and/or because they were neither positive nor negative evaluations.

| **Themes** | | **Survey codes^1^** | **Interview codes^1^** |
| --- | --- | --- | --- |
| Bohren et al. themes | | | |
|  | Physical abuse/sexual abuse | Assault | - |
|  | Verbal abuse | Insults, intimidation, causing guilt, threats | Threats |
|  | Failure to meet professional standards of care | Lack of consent, lack of consent but ok, pressure, vaginal examinations, CTG, fundal pressure during delivery, fundal pressure for placenta expulsion, membrane stripping, inadequate pain relief, lack of support | Pressure, lack of informed choice, lack of informed consent, feeling abandoned, neglect, waiting, painful vaginal examinations, lack of privacy |
|  | Poor rapport between women and providers | Not taken seriously, not taken seriously but ok, lack of information, degradation, unprofessional conduct, lack of collaboration between HCP, rough treatment, language issues, restriction of freedom, lack of respect for wishes/feelings/needs | Lack of interpersonal fit, unprofessional behaviour, degradation, not taken seriously, lack of transparency, conflicting information, lack of information, feeling passive, uncomfortable birthing position, lack of individualised care, lack of respect for needs, discouragement, medication in lieu of support, lack of empathy |
|  | Health system conditions and constraints | Inadequate facility/resources | Hectic atmosphere, facility overstretched, staff stressed, staff out of their depth, shift change, no childbirth debriefing |
| Interview themes | | | |
|  | Informal coercion |  | Threats, pressure, lack of informed choice, lack of informed consent, lack of privacy, not taken seriously, painful vaginal examinations |
|  | Risk factors for mistreatment |  | Inadequate antenatal classes, feeling insecure, helplessness, lack of coping, no longer being able to think, prolonged labour, partner persuades woman, trust in HCP |

| Interview themes, continued | | | |
| --- | --- | --- | --- |
|  | Consequences of mistreatment |  | Negative birth experience, loss of control, feeling at HCP’s mercy, panic, assertiveness, opposition, negotiating a compromise, accepting HCP’s opinion, giving in, guilt, self-deprecation, self-doubt, thinking about the birth, «what if», need for clarification, attitude change, frustration, sadness, regret, thoughts concerning future birth, avoidance, helpful childbirth debriefing, processing, extenuation |
|  | Good examples of care |  | One-on-one care, empathetic care, respecting boundaries, informed choice, autonomy, transparent communication, providing time |
|  | What’s needed |  | What’s needed |
| Miscellaneous^2^ | | | |
|  | Childbirth debriefing | Not helpful, not offered, HCP not available, transfer, extra charge, timing, not yet |  |
|  |  |  | Fear of caesarean section, fear of vaginal birth, exceptional situation, CTG negative, straightforward communication, involving partner, social norms, separation from child, preparation |
